# Excess Mortality in the United States During the First Three Months of the COVID-19 Pandemic

**DOI:** 10.1101/2020.05.04.20090324

**Authors:** R. Rivera, J. E. Rosenbaum, W. Quispe

## Abstract

Deaths are frequently under-estimated during emergencies, times when accurate mortality estimates are crucial for emergency response. This study estimates excess all-cause, pneumonia, and influenza mortality during the COVID-19 pandemic using the September 11, 2020 release of weekly mortality data from the United States (U.S.) Mortality Surveillance System (MSS) from September 27, 2015 to May 9, 2020, using semiparametric and conventional time-series models in 13 states with high reported COVID-19 deaths and apparently complete mortality data: California, Colorado, Connecticut, Florida, Illinois, Indiana, Louisiana, Massachusetts, Michigan, New Jersey, New York, Pennsylvania, and Washington. We estimated greater excess mortality than official COVID-19 mortality in the U.S. (excess mortality 95% confidence interval (CI) (100013, 127501) vs. 78834 COVID-19 deaths) and 9 states: California (excess mortality 95% CI (3338, 6344) vs. 2849 COVID-19 deaths); Connecticut (excess mortality 95% CI (3095, 3952) vs. 2932 COVID-19 deaths); Illinois (95% CI (4646, 6111) vs. 3525 COVID-19 deaths); Louisiana (excess mortality 95% CI (2341, 3183) vs. 2267 COVID-19 deaths); Massachusetts (95% CI (5562, 7201) vs. 5050 COVID-19 deaths); New Jersey (95% CI (13170, 16058) vs. 10465 COVID-19 deaths); New York (95% CI (32538, 39960) vs. 26584 COVID-19 deaths); and Pennsylvania (95% CI (5125, 6560) vs. 3793 COVID-19 deaths). Conventional model results were consistent with semiparametric results but less precise. Significant excess pneumonia deaths were also found for all locations and we estimated hundreds of excess influenza deaths in New York.

We find that official COVID-19 mortality substantially understates actual mortality, excess deaths cannot be explained entirely by official COVID-19 death counts. Mortality reporting lags appeared to worsen during the pandemic, when timeliness in surveillance systems was most crucial for improving pandemic response.

## Introduction

The number of Coronavirus Disease 2019 (COVID-19) deaths may be under-reported, and COVID-19 may be indirectly responsible for additional deaths. The Centers for Disease Control and Prevention issues guidelines to determine cause of deaths, but underestimating the death toll of natural disasters, heatwaves, influenza, and other emergencies is common. In some cases, the underestimates can be extreme: chikungunya was officially associated with only 31 deaths during a 2014-15 epidemic in Puerto Rico, but time series analysis estimated excess mortality of 1310 deaths [1]. Hurricane Maria’s mortality was officially only 64 deaths, but a 95% confidence interval for estimated excess mortality was between 1,069 and 1,568. [2] Deaths directly due to the COVID-19 pandemic may be underestimated due to under-diagnosis [3], insufficient postmortem Severe Acute Respiratory Syndrome Coronavirus 2 (SARS-CoV-2) tests, not seeking healthcare [4], and ascertainment bias. Deaths indirectly due to an emergency are also common, due to an overloaded health system [5] or lack of healthcare access for routine care: during the 2014 West Africa Ebola epidemic, lack of routine care for malaria, HIV/AIDS, and tuberculosis led to an estimated 10,600 additional deaths in the area.[6] Health emergencies may also lead to indirect deaths from economic, social, and emotional stress [7] and crowded emergency departments.[8]

In this study, we estimate excess all-cause, pneumonia, and influenza mortality during the COVID-19 pandemic, which includes deaths both directly and indirectly related to COVID-19. Directly related COVID-19 deaths include deaths in patients who have undetected SARS-CoV-2 due to false negative tests [9] not seeking healthcare [4], or being turned away from the emergency department due to emergency department crowding [8]. Testing and forensic staff shortfalls lead to lack of postmortem testing. Indirect deaths are deaths not due to COVID-19 and may include deaths among patients due to emergency department crowding [8]; avoidance of hospitals due to fear of the virus; or avoidance of healthcare due to the accompanying economic recession, such as loss of employment or income, or loss of health insurance coverage.[10]

## Methods

### Data and measures

Weekly all-cause, pneumonia, and influenza mortality for each state from September 27, 2015 (week 40) to May 9, 2020 (week 19) were obtained from the National Center for Health Statistics (NCHS) Mortality Surveillance System (MSS) data release on September 11, 2020. The MSS presents weekly death certificate counts, without regard to whether deaths were classified as related to COVID-19. Based on ICD-10 multiple cause of death codes, pneumonia and influenza deaths were also made available. The timeliness in death certificate reporting varies by region, state, and cause of death.[11] We use pneumonia and influenza mortalities because COVID-19 deaths could be misclassified as pneumonia or influenza. The NCHS divides New York State into two jurisdictions: New York City (NYC) and non-NYC New York State, and we leave them separate for plots and combine them for statistical models.

We identified 13 jurisdictions within this data that had high numbers of reported COVID- 19 deaths through May 9, 2020. The states were: California, Colorado, Connecticut Florida, Illinois, Indiana, Louisiana, Massachusetts, Michigan, New Jersey, New York, Pennsylvania, and Washington.

Population estimates were from Vintage 2019 Census yearly estimates for July 1 of each year 2010–19, which were used to obtain weekly population estimates.[12]

We obtained COVID-19 mortality counts through 5/9/2020 from the COVID-19 Tracking Project, the New York Times, and the Centers for Disease Control Provisional Deaths. Usually, CDC provisional counts were the highest estimate [13, 14, 15]. These three mortality counts differ, so we used the higher number in all cases to be conservative with respect to the null hypothesis that excess mortality can be explained entirely by official COVID-19 death counts: that is, the lower bound of excess mortality is below official COVID-19 deaths. To assess whether the COVID-19 pandemic was associated with reduced emergency department (ED) utilization not for COVID-19, we identified 3 of the United States top 5 causes of death that present with acute symptoms that require immediate treatment, for which the choice not to seek healthcare may result in death: heart disease, chronic lower respiratory diseases, and cerebrovascular disease. Among the 59 National Syndromic Surveillance Program (NSSP) jurisdictions, we were only able to obtain daily ED visits in New York City for asthma symptoms. We obtained daily counts of asthma ED visits, age group (5–17, 18–64, 65+), borough, and date from the New York City Department of Health and Mental Hygiene’s EpiQuery website from January 1, 2016 to May 9, 2020. This study is an analysis of publicly available data in broad categories such that individuals cannot be identified, so it is not human subjects research and is exempt from requiring human subjects board review.

### Statistical Analysis

We construct two models to capture the temporal behavior of death certificate data to estimate excess mortality during the COVID-19 pandemic: a semiparametric model and a conventional model estimating the difference in current death totals starting from the beginning of the pandemic and the projected deaths under normal conditions [16]. In what follows, we briefly describe each excess deaths model. See Appendix for further details.

### Semiparametric model

We use a semiparametric model to capture the temporal behavior of mortality while measuring how the pandemic alters the ‘normal’ mortality pattern. This model was successfully deployed to estimate excess deaths due to Hurricane Maria in Puerto Rico [2].

Specifically, we define a general additive model with the following covariates: the natural logarithm of population as an offset, a smooth function of week of the year, year category, and a binary variable coded as 1 for dates on or after the start of the pandemic and 0 prior to the starting point; its coefficient presents the possibility that the mean death rate has changed after the start of the pandemic at some location. Week of the year is modeled non-parametrically with a penalized cyclic cubic regression spline function to capture seasonal mortality variations [15]. Preliminary analysis indicated overdispersion, so we used a quasi-Poisson model to estimate the dispersion parameter [17]. We estimated coefficients by a penalized likelihood maximization approach, where the smoothing penalty parameters are determined by restricted maximum likelihood. The residuals of the fitted model did not present remaining temporal dependence.

To estimate cumulative excess deaths, we sum the coefficients of the indicator for the start of the pandemic. Approximate simulations from the Bayesian posterior density are performed to obtain 95% credibility intervals, which we refer to as confidence intervals throughout.

For each state, we determine excess deaths due to the COVID-19 pandemic from a starting point through May 9, the date of the most recent complete mortality data at the time of our analysis. The starting point was the date after the most recent inflection point in all-cause mortality, suggesting the onset of the COVID-19 pandemic, chosen to balance concerns of deaths prior to the official first cases and the sensitivity of the model to detect small excess mortality in the limited available data, exacerbated by the provisional counts being lower than actual deaths: February 29 for Washington State, March 28 for Florida, Indiana, and Massachusetts, and March 21 for the remaining 9 states and for the United States as a whole. This semiparametric model can incorporate population displacement [2], but we are not aware of significant displacement during the pandemic. Specifically, New York City was uniquely affected during this first wave of infections, with a public perception of widespread community infections acquired in a crowded metropolitan area that relies on public transportation. Analysis of data from smart phones finds that about 400,000 New York City residents, 5% of the population of New York City, left the city between March 1 and May 1, 2020, and departures were characterized by substantial wealth and race disparities [18]. NYC residents dispersed to locations around the country, with most locations receiving less than 4000 people from this total, a negligible addition (<1%) to the state populations; locations receiving more than 4000 people included non-NYC New York State, Pennsylvania, New Jersey, Connecticut, and Florida [18].

### A conventional excess deaths analysis

We also estimate excess mortality using a conventional excess mortality method: we fit a quasi-Poisson semiparametric model as above until February 1, 2020. The deaths from February 8, 2020 forward should follow a Poisson distribution with some expected rate; the maximum likelihood estimator of such rate is the mean weekly deaths during this period. Weekly deaths are approximately normally distributed with a variance that accounts for overdispersion according to the scale parameter in the fitted model. From this distribution, we simulate 10,000 weekly deaths and subtract the results from the posterior distributions of the fitted results. The 95% confidence interval is the range between the 2.5% and 97.5% percentiles of all excess deaths.

The models were fit using version 1.8-28 of the *mgcv* package in R 4.0.2 [19, 20]. All code and data have been made publicly available (https://github.com/bakuninpr/COVID-Excess-Deaths-US). The statistical analyses were conducted between April and September 2020.

### Assessing reduction in emergency department utilization

To evaluate whether there was a reduction in emergency department visits during the period from March 21 to May 9, we fit a quasi-Poisson regression model for daily number of asthma-related emergency department visits in New York City syndromic surveillance data, controlling for borough, age group, and date, with a binary variable for date after 3/21/20; this model estimated a dispersion parameter of 1.95.

## Results

Provisional death counts increase with each data release, especially for recent weeks. The variation on reporting timeliness by state hinders excess death assessment for the United States. Figure 1 shows all-cause mortality counts for successive data releases. Successive releases of weekly mortality counts ‘blanket’ each previous release, with gaps between the lines representing differences in each data release. During the pandemic, large gaps between successive data releases during periods of higher deaths suggests that mortality reporting lags are larger during periods of elevated deaths: the discrepancy between the number of all-cause deaths for the last week of the April 1^st^ data release and the corresponding entry of the June 12 release is 18%. In contrast, the discrepancy between the number of all-cause deaths for the last week of the May 29 data release and the corresponding entry of the June 12 release is 75%. The effects on mortality of the COVID-19 pandemic is clearly seen in Figure 2, although the effect has substantial local variation. All states have excess pneumonia mortality (Figure 3), and New York City shows hundreds of excess influenza deaths over several weeks (Figure 4).

**Figure 1:**
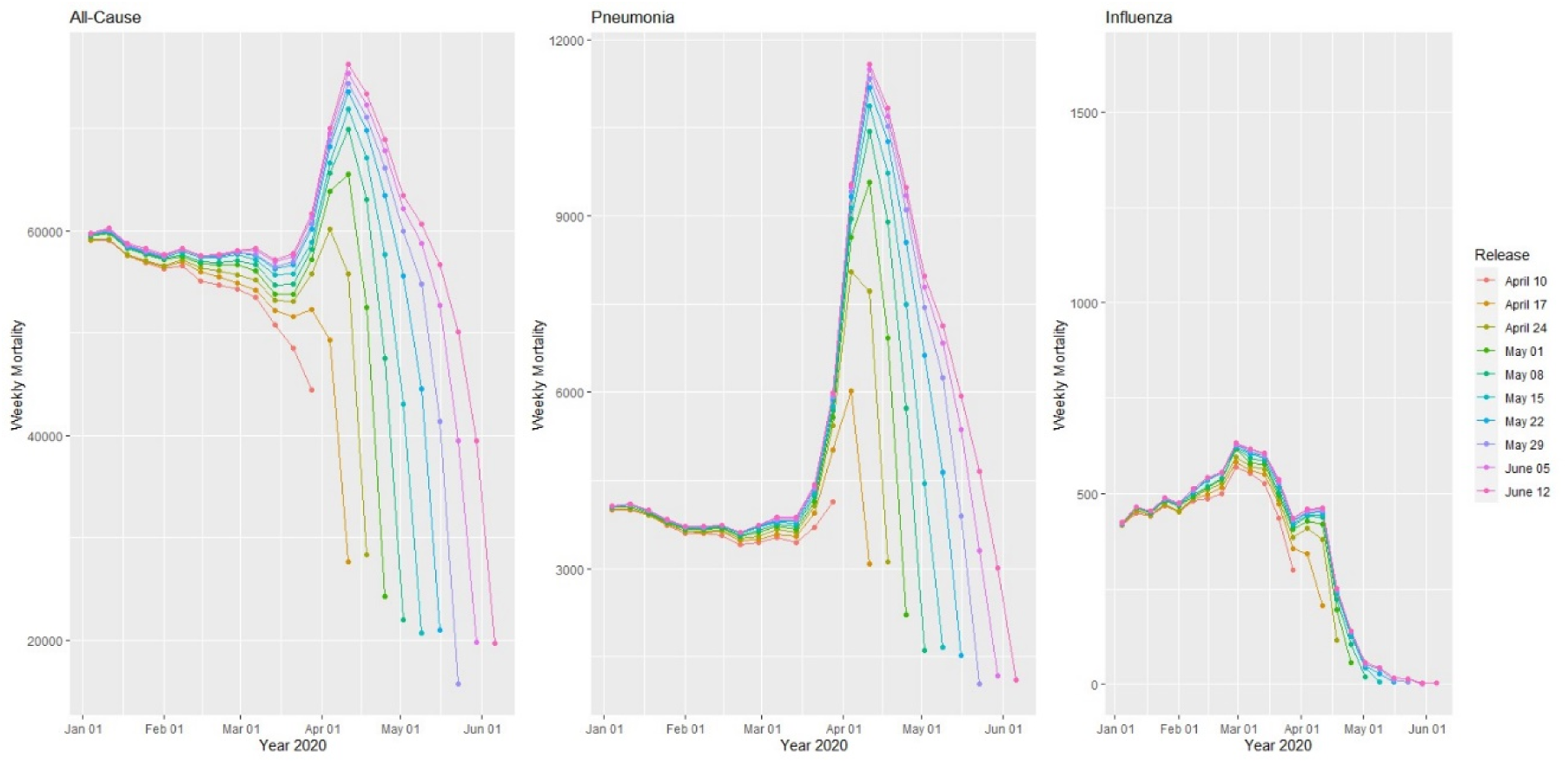
Provisional weekly all-cause, pneumonia and influenza mortality counts for the United States from weekly data releases April 10–June 12, 2020.

**Figure 2:**
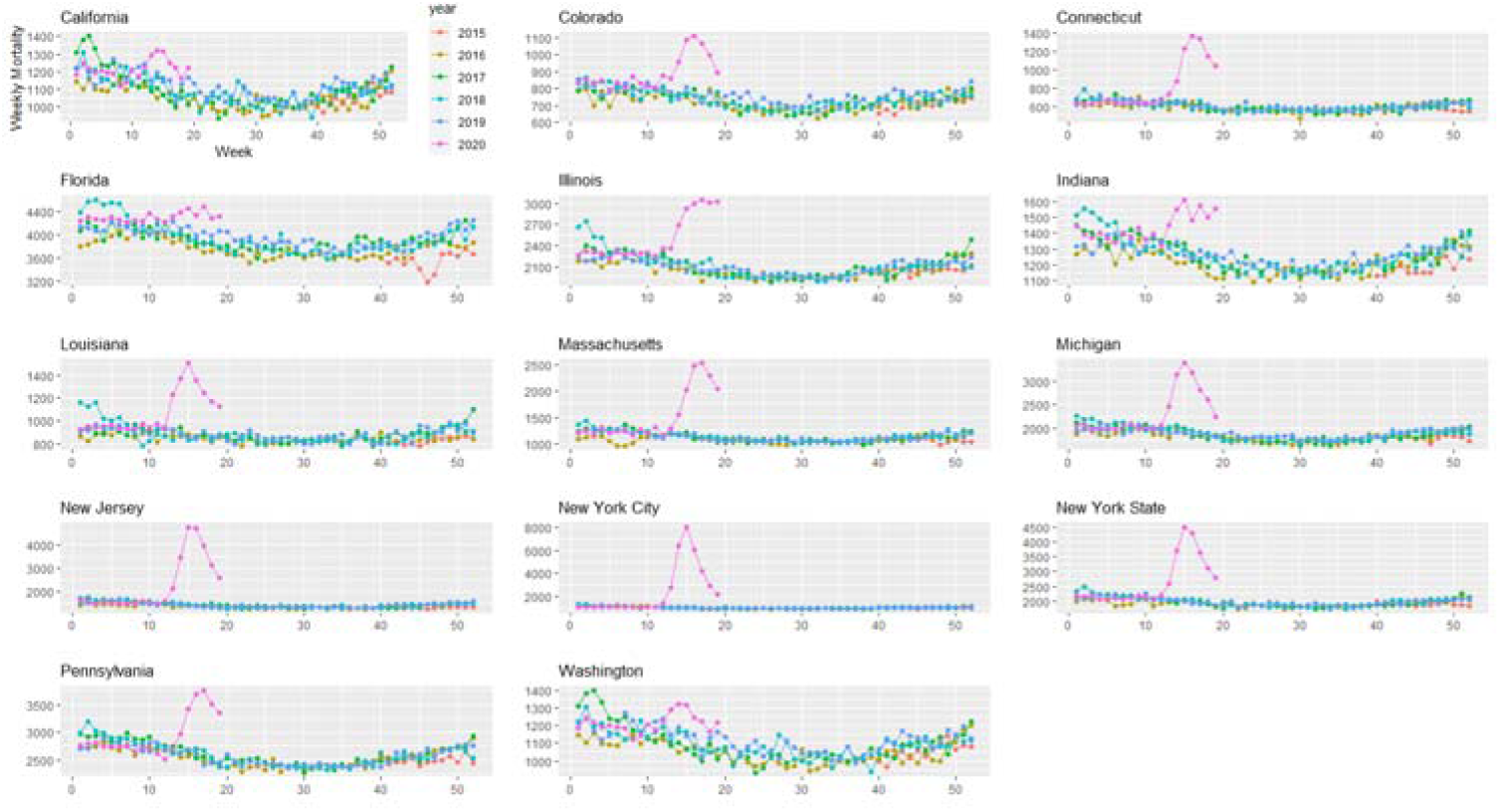
Weekly all-cause mortality grouped by year and state starting on week 40 of 2015 until 5/9/2020.

**Figure 3:**
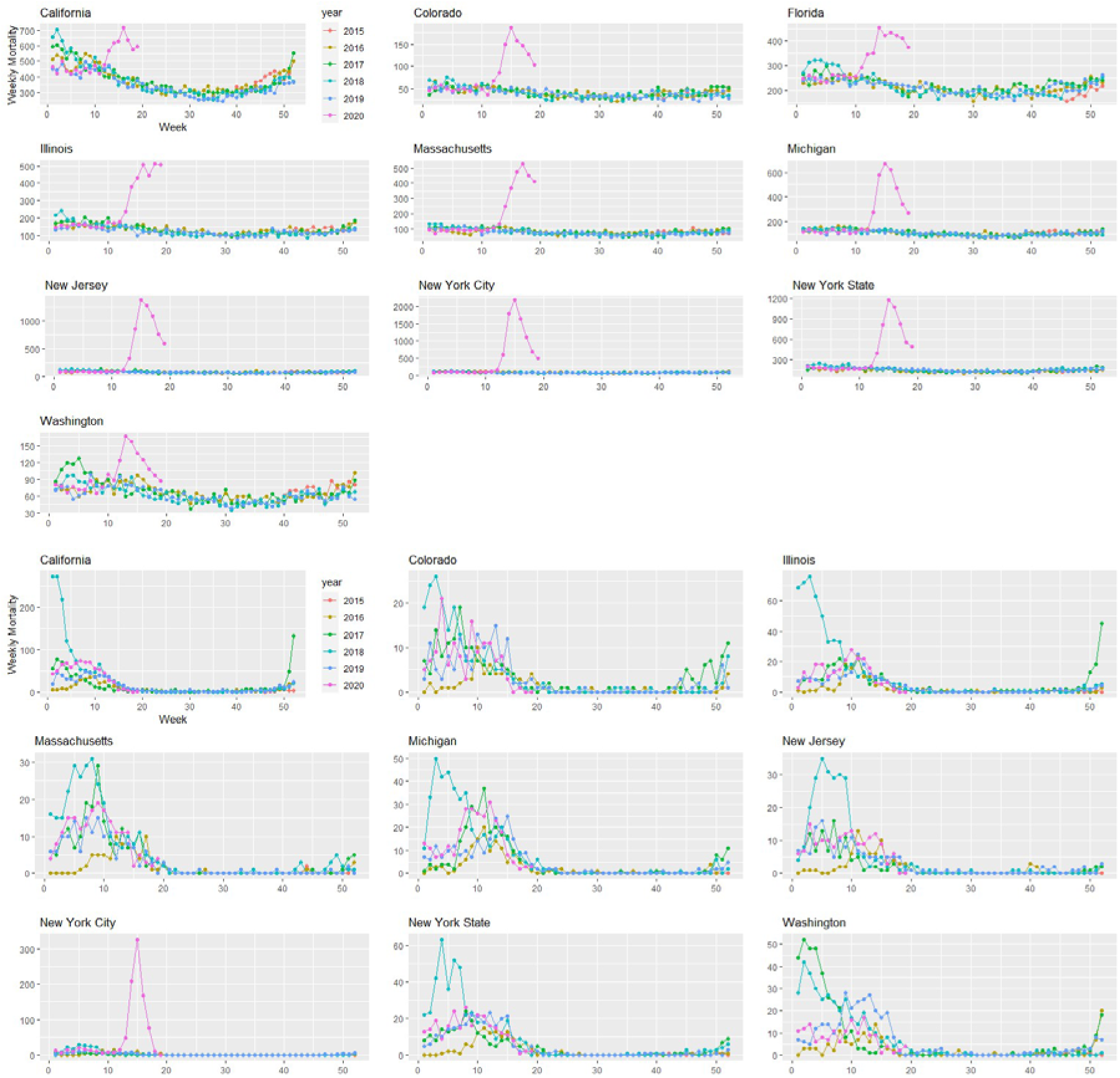
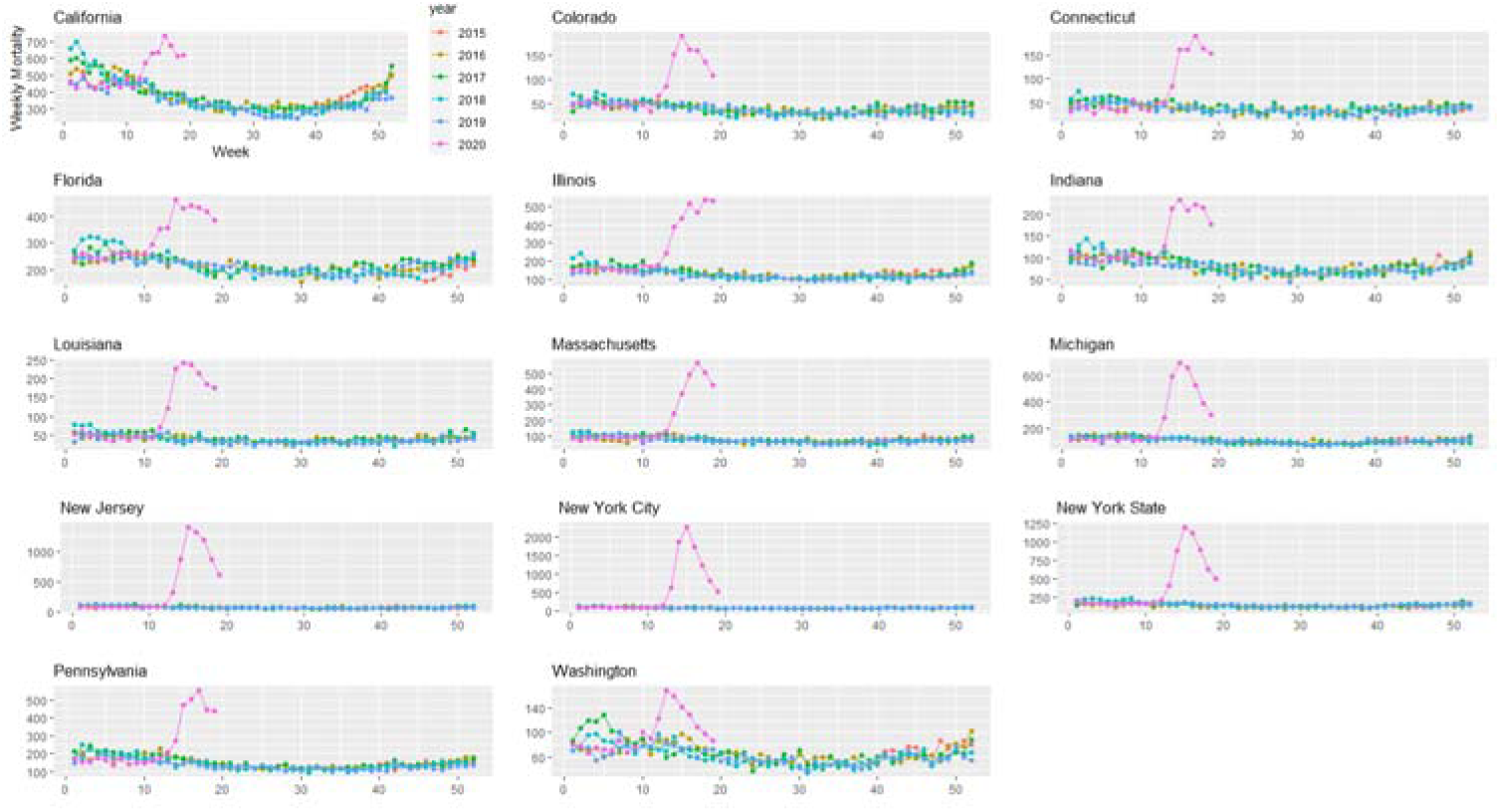
Weekly pneumonia mortality grouped by year and state starting on week 40 of 2015 until 5/9/2020.

**Figure 4:**
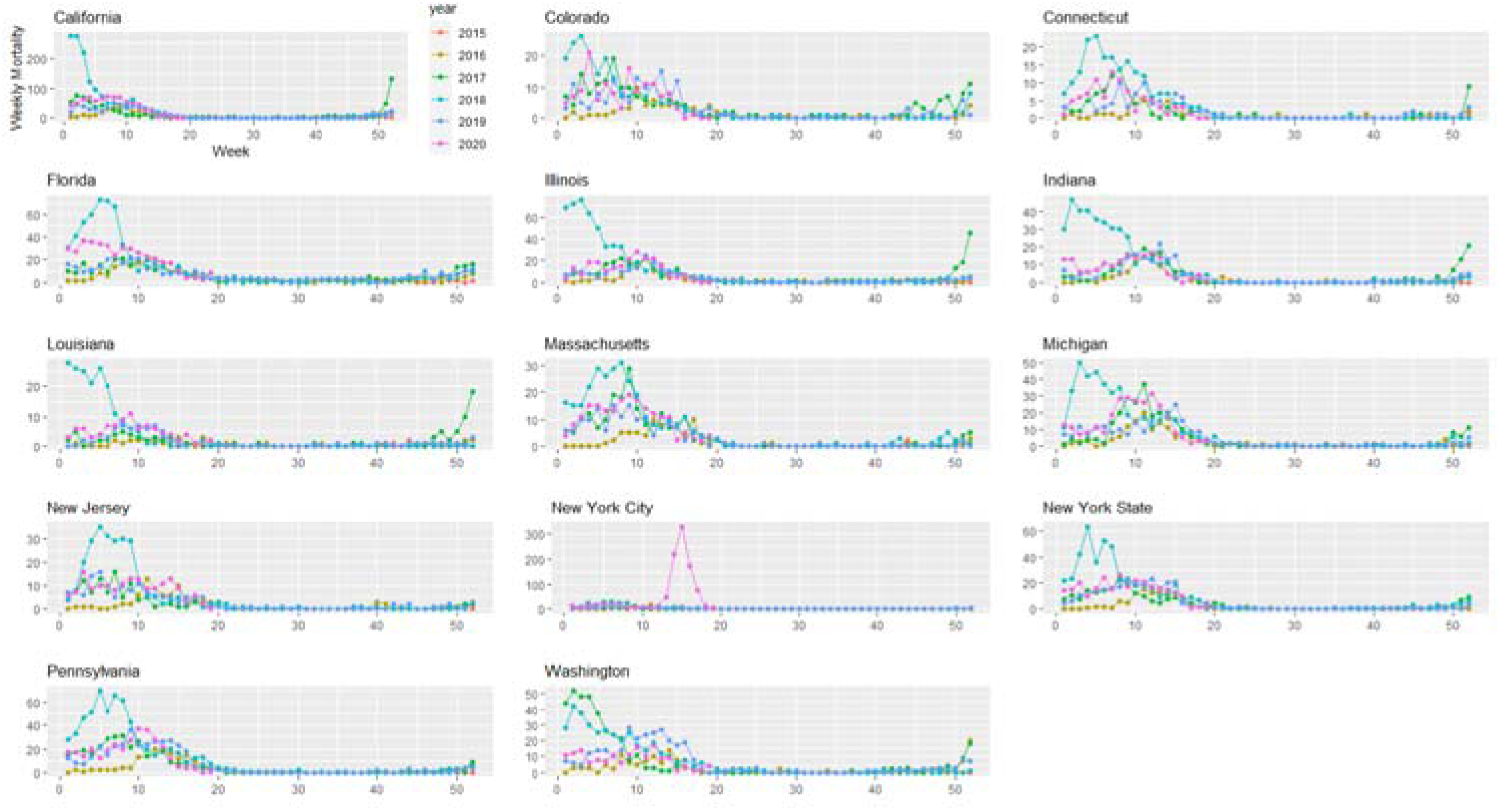
Weekly influenza mortality grouped by year and state starting on week 40 of 2015 until 5/9/2020.

In the United States, we observe greater all-cause and pneumonia deaths (Figure 5). Using the semiparametric model, between March 21, 2020 and May 9, 2020 we are 95% confident that all-cause excess deaths in the United States were between 100013 and 127501, compared with 78834 reported COVID-19 deaths, which is at least 21179 more deaths than official COVID-19 deaths for the same time period. Pneumonia excess deaths were between 40066 and 47391. Using the conventional method, we found a 95% confidence interval of 106940 and 143478 for all-cause excess deaths in the United States, and 41613 and 47841 for pneumonia deaths.

**Figure 5:**
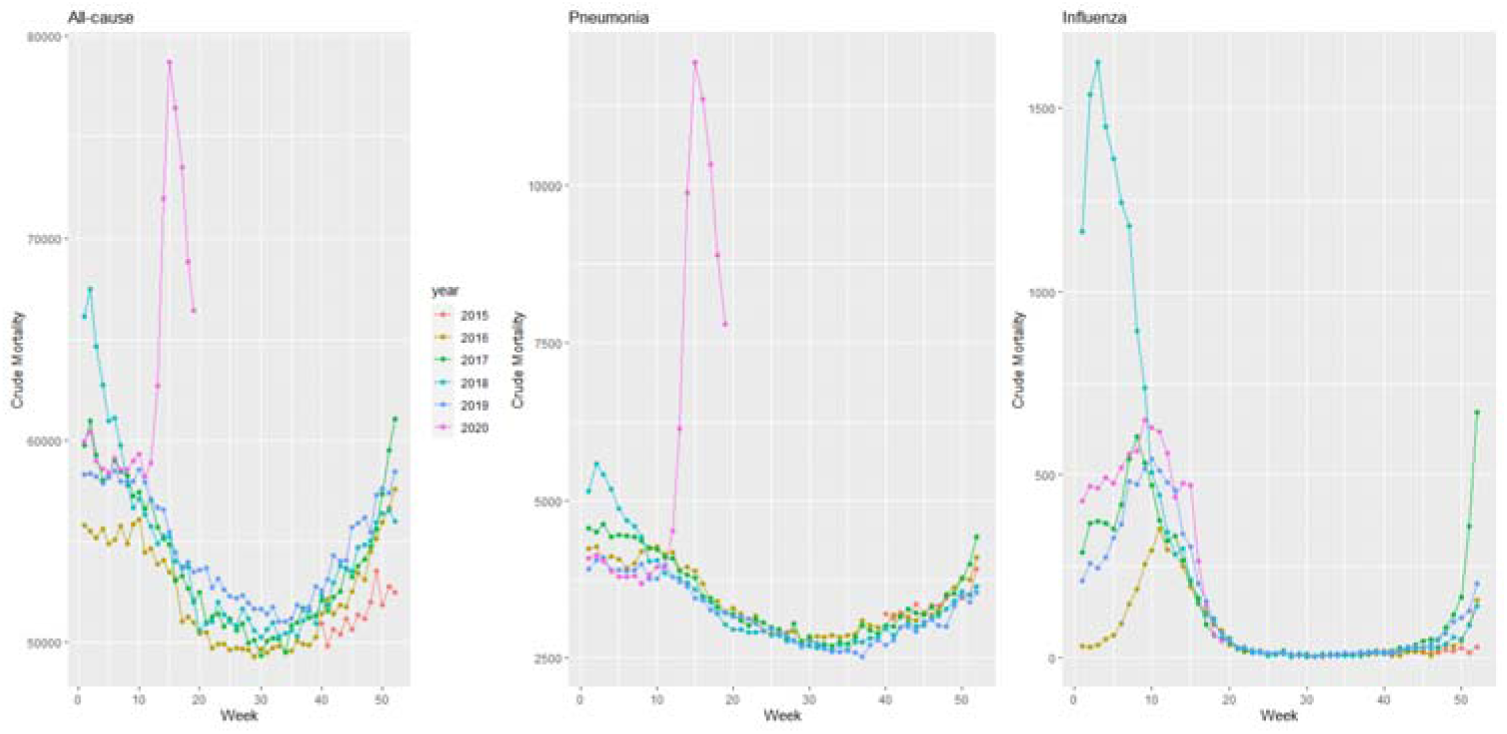
Weekly mortality in the United States by year starting on week 40 of 2015 until 5/9/2020.

For each state Table 1 presents 95% confidence intervals for all-cause, pneumonia, and influenza excess deaths. We estimated greater excess mortality than COVID-19 deaths in 10 of 13 states: Florida, Indiana, and Washington present significant all-cause excess mortality but there was no evidence of them exceeding official COVID-19 death counts. However, relative to expected numbers of pneumonia and influenza deaths, we observe excess pneumonia deaths in all states studied, and excess influenza deaths in New York while excess influenza deaths in New Jersey are less clear (Table 1). We evaluated influenza mortality in the District of Columbia, the other urban-only area, but influenza deaths were not higher than usual (not shown), in contrast to New York City.

**Table 1:** Excess all-cause, pneumonia, and influenza mortality 95% confidence intervals from 2 models, from states with the largest reported COVID-19 mortality data as of 5/9/2020, and with official COVID-19 toll. All data used included mortality until 5/9/2020 included in the 9/04/2020 data release. The semiparametric method uses a starting date, but the conventional method does not. The table presents prediction intervals using the conventional method, although we refer to confidence intervals throughout for consistency.

| Jurisdiction | Start Date | COVID-19 | All-cause Excess Deaths |  | Pneumonia Excess Deaths |  | Influenza Excess Deaths |  |
| --- | --- | --- | --- | --- | --- | --- | --- | --- |
|  |  | Deaths | Semiparametric | Conventional | Semiparametric | Conventional | Semiparametric | Conventional |
| California | 3/21/2020 | 2849 | (3338, 6344) | (3638, 9058) | (1729, 2370) | (1969, 2900) | (-75, 52) | (120, 359) |
| Colorado | 3/21/2020 | 1130 | (1175, 1730) | (960, 1882) | (620, 803) | (557, 807) | (-26, 15) | (-61, 36) |
| Connecticut | 3/21/2020 | 2932 | (3095, 3952) | (3092, 4008) | (651, 844) | (638, 864) | (-20, 5) | (-63, 18) |
| Florida | 3/28/2020 | 1840 | (1271, 2856) | (392, 3934) | (1100, 1439) | (1373, 1966) | (-39, 21) | (-200, 7) |
| Illinois | 3/21/2020 | 3525 | (4646, 6111) | (5111, 7357) | (1974, 2422) | (2088, 2566) | (-20, 44) | (8, 129) |
| Indiana | 3/28/2020 | 1490 | (1400, 2078) | (1198, 2613) | (679, 882) | (588, 941) | (-28, 14) | (-60, 66) |
| Louisiana | 3/21/2020 | 2267 | (2341, 3183) | (2522, 3834) | (1042, 1263) | (970, 1288) | (-3, 27) | (14, 68) |
| Massachusetts | 3/28/2020 | 5050 | (5562, 7201) | (5516, 7152) | (2044, 2456) | (2012, 2439) | (-100, 39) | (-111, 34) |
| Michigan | 3/21/2020 | 5036 | (5581, 7171) | (5514, 7474) | (2386, 2926) | (2458, 2910) | (-52, 28) | (-36, 138) |
| New Jersey | 3/21/2020 | 10465 | (13170, 16058) | (13834, 15567) | (5550, 6539) | (5879, 6375) | (2, 46) | (-46, 61) |
| New York | 3/21/2020 | 26584 | (32538, 39960) | (35632, 38802) | (12016, 14310) | (12860, 13623) | (694, 911) | (640, 930) |
| Pennsylvania | 3/21/2020 | 3793 | (5125, 6560) | (4653, 6809) | (1757, 2135) | (1686, 2135) | (-65, -8) | (-96, 90) |
| Washington | 2/29/2020 | 925 | (559, 1633) | (421, 1796) | (358, 623) | (359, 674) | (-3, 72) | (-20, 78) |
| United States | 3/21/2020 | 78834 | (100013, 127501) | (106940, 143478) | (40066, 47391) | (41613, 47841) | (15, 1385) | (-33, 2896) |

With the conventional model, we estimate more excess all-cause deaths than reported COVID-19 cases except in Colorado, Florida, Indiana, and Washington (Table 1); and once more excess pneumonia deaths are observed for all states. These results are also consistent with the excess mortality results for the earlier data release covering through May 9, 2020 (not shown).

The results of the semiparametric and conventional models are generally compatible. But since our conventional model also reflects random fluctuations in mortality, prediction intervals are obtained, which naturally present larger variation than confidence intervals. Treating the deaths as fixed [16] would lead to intervals that are misleadingly short on width. The larger uncertainty in the conventional model makes it harder to interpret excess deaths.

Using the quasi-Poisson model with the outcome daily emergency department visits for asthma syndrome in NYC, we found that during this period, asthma visits were 64% lower than expected, a substantial drop(IRR = 0.36, 95% CI (0.34, 0.37)).

## Discussion

We find substantial excess all-cause mortality that exceeds the number of documented COVID-19 deaths in most of the 13 states evaluated: California, Colorado, Connecticut, Illinois, Louisiana, Massachusetts, Michigan, New Jersey, New York, and Pennsylvania. Up until May 9^th^, we estimate over 100,000 excess deaths in the U.S. due to the COVID-19 pandemic. Mortality underestimation may reduce the public’s willingness to adhere to costly and stressful non-pharmaceutical interventions, such as governors’ orders to stay at home, wear masks, and engage in social distancing.

### Mechanisms for excess mortality

The CDC has established guidelines for certifying COVID-19 deaths [21] and whether to collect postmortem specimens for SARS-CoV-2 testing [22]. We propose the following potential mechanisms for underestimation of pandemic death toll: underdiagnosis of COVID-19 due to low availability of SARS-CoV-2 tests; indirect deaths from not seeking care for emergent non-COVID-19 conditions; not seeking care for what appeared to be non-severe COVID-19 and then experiencing sudden declines characteristic of COVID-19; or needing treatment for COVID-19 or other ailments and being turned away from emergency departments due to crowding, and subjective interpretations of guidelines. COVID-19 test access has been quantified as the percent of SARS-CoV-2 polymerase chain reaction (PCR) tests that are positive; the percent of PCR tests that are positive decreased during this period, suggesting increased test access [24]. However, substantial heterogeneity in test availability across states [13] means that excess mortality may not decrease substantially unless test availability increases in high-prevalence states.

Some excess mortality may include indirect deaths from emergent non-COVID-19 conditions due to delaying healthcare, due to fear of becoming infected with SARS-CoV-2 during the COVID-19 pandemic. Other research suggests that patients with heart attacks and stroke delayed seeking care due to the COVID-19 pandemic [25]. Our research suggests some deaths could include deaths from chronic lower respiratory diseases, such as deaths due to not seeking care for asthma syndrome. In this research, we were not able to access emergency department visit data for acute coronary syndromes, only asthma syndrome ED visits in NYC. When the complete 2020 mortality data are released, we would expect more deaths at home coded as chronic lower respiratory diseases, cardiovascular disease, and cerebrovascular diseases than usual; normally, these are three of the top five causes of death in the US, so reduced care-seeking could contribute substantially to excess mortality [26].

Patients who suspect COVID-19 may not seek healthcare or may be turned away from emergency departments: these patients would not have SARS CoV-2 test results, so they would not be coded as COVID-19 deaths. Many patients with influenza-like illness (ILI) never seek healthcare, including patients likely to have severe effects: about 45% of patients with heart disease and 52% with COPD delay at least 3 days [27], and during a flu pandemic care-seeking increases by only about 10 percentage points [4]. Dyspnea/breathlessness predicts healthcare seeking for ILIs [28]. However, most severe or fatal COVID-19 cases do not present with dyspnea [29], and lung damage can be substantial even without dyspnea [30]. Sudden health declines have been observed in COVID-19 inpatient populations: patients decline in minutes from ambulatory/conversant to unresponsive and requiring resuscitation [31], and such sudden health declines could also occur in outpatient populations.

Some but not all of the excess COVID-19 mortality is captured by pneumonia excess mortality, but some sudden deaths seem to occur among patients without apparent pneumonia [32]. These deaths may be due to cardiac injury [33], kidney or liver injury [34], or hypercoagulability [35]. Further, our results suggest that excess deaths for cardiovascular, cerebrovascular, kidney or liver failure without coding for COVID-19 may be apparent when these data become available.

### Advantages of the semiparametric model

The CDC estimates and publishes excess mortality counts during the COVID-19 pandemic, but without quantifying uncertainty like our methods do. It can be shown that under a mixed model representation, the semiparametric model estimates are the best linear unbiased predictors [36]. Moreover, the intervals from our two methods always overlap. Yet the semiparametric method yields more precise confidence intervals than a conventional approach, which must estimate prediction intervals. Prediction intervals are wider than confidence intervals because they account for uncertainty in post-pandemic deaths. That is, for week 19 of 2020, the conventional model estimates excess deaths as the difference between the observed deaths, and what pre-pandemic projected forward model expects the number of deaths to be. Those observed deaths in week 19 are subject to random variation in mortality which the interval must account for. In contrast, the semiparametric model estimates excess deaths as the difference of two expected values: expected mortality with a pandemic period indicator and expected mortality without a pandemic period indicator. Intervals of parameters are always narrower than for random variables. Wider intervals hinder interpretation of excess deaths, so we focus on the results from the semiparametric model. The semiparametric model is also less affected by under-reporting during the pre-pandemic period than the conventional approach.

### Misclassification of excess deaths

We quantify higher pneumonia mortality than expected in all 13 states and higher influenza deaths primarily in New York. For New Jersey, even when adjusting for COVID-19 official deaths, pneumonia and influenza mortality, excess deaths are still significant, which raises two potential explanations: COVID-19 deaths may have been misclassified or New Jersey may have had more indirect deaths than other states. With only limited cause of death data, our analysis is unable to distinguish between misclassified COVID-19 deaths and indirect deaths.

Excess influenza deaths in New York City appear to be misclassified COVID-19 deaths. If NYC’s COVID-19 emergency declaration led people with seasonal influenza not to seek care, we would expect to see a sharp reduction in the number of positive influenza cases around the date of the emergency declaration. However, the number of positive influenza tests in New York City decreased steadily throughout March 2020 with no steep drop around the date of the COVID-19 emergency declaration [23]. Because seasonal influenza seemed to taper off during March 2020, while influenza deaths in NYC increased until April 11th. This suggests that many excess influenza deaths were misclassified COVID-19 deaths, rather than a resurgence of undiagnosed influenza. The apparent misclassification of COVID-19 deaths as influenza in New York City does not appear attributable to urbanicity because we observed no excess influenza mortality in the District of Columbia, the other urban-only area The likely misclassification of COVID-19 deaths as influenza may be due to heterogeneity in cause of death determination and/or COVID-19 presentations between New York City and the states examined. Alternatively, the misclassification may occur in many jurisdictions, but it is more detectable in NYC due to the large number of deaths.

### Mortality displacement hypothesis

Excess mortality during heatwaves and influenza pandemics often shows mortality displacement (or harvesting effect), where subsequent mortality declines [37]. Longer observation periods could detect mortality displacement, but inconsistent non-pharmaceutical intervention policies across the United States may cause elevated mortality due to COVID-19 to persist over longer durations than during typical influenza pandemics.

## Strengths and Limitations

Our hypothesized mechanisms for excess mortality are based on existing research, but the available mortality data are insufficient to test these hypotheses. For example, deaths at home are not published weekly or systematically. The available U.S. data includes data aggregated over all states stratified by age (0–17, 18–64, 65+) and region; however, these data do not allow identification of all-cause mortality trends by region or age because some states’ data are incomplete, even by the available completeness measure, which understates completeness during periods of elevated mortality.

Our analysis examines excess deaths associated with the COVID-19 pandemic. It is likely that the longer the period since the beginning of the pandemic, the more indirect causes of death are included in our excess deaths estimate. The vast majority of excess mortality is not attributable to social distancing or the pandemic-induced recession because recessions are generally associated with lower mortality [38]. Furthermore, the statistical methods used can estimate excess mortality in regions with considerable increases in fatalities. In regions with very small number of increased deaths, our methods may not be able to detect the effects of the pandemic because the confidence intervals would be too wide.

We can evaluate whether emergency department visits for asthma in New York City decreased during the COVID-19 pandemic but not whether the decrease in asthma ED visits increased mortality because chronic lower respiratory mortality data from this period are not currently available. We were unable to obtain data from the other 58 NSSP jurisdictions, or for acute coronary syndromes that could suggest stroke or heart attack.

### Implications for policy and practice

Research can identify populations at risk for mortality that may not be included in the official COVID-19 counts, and the overlap with groups affected by other health disparities, so resources can be allocated to the most affected communities. Excess mortality may be reduced by prioritizing research, including random sample testing of people without symptoms and postmortem tests [39]. To avoid ascertainment bias, random sample postmortem testing can identify atypical disease presentations that may be under-recognized in clinical settings. In the face of testing limitations, postmortem tests may be viewed as expendable, but postmortem testing can improve patient care by identifying gaps in diagnosis and treatment.

To identify under-use of healthcare for cardiovascular and cerebrovascular conditions, National Surveillance System Program data should include these indications and current data should be available for all NSSP locations. New York City’s system suggests that this change to make data more quickly available is feasible. NSSP jurisdictions are given substantial discretion over which syndromes are included and may consider centralized standards impractical, but such standards would make data useful nationwide, in addition to locally.

Future federal pandemic planning should include upgrades to state vital statistics infrastructure, so that all states can report deaths in a timely fashion. As suggested after earlier pandemics [40], pandemic planning should identify how to release more detailed data so that research can discriminate between mechanisms of excess mortality, especially for high-vulnerability jurisdictions. Research using detailed data will find some spurious findings, including cases who died with the virus but not of the virus, but more detailed data will save lives by identifying vulnerable communities to allocate resources to, as well as atypical symptoms of the disease.

## Conclusions

Excess all-cause mortality exceeding the number of reported COVID-19 deaths is evident in the United States and in many states during the first three months of the pandemic. Greater test availability, including postmortem tests, can yield more accurate mortality counts and case-fatality ratios, and increase the public’s willingness to adhere to non-pharmaceutical interventions to reduce transmission.

## Data Availability

Data comes from National Center for Health Statistics and is available to the public: https://gis.cdc.gov/grasp/fluview/mortality.html

https://github.com/bakuninpr/COVID-Excess-Deaths-US

## Abbreviations

CI: confidence intervals
COVID-19: Coronavirus Disease 2019
ED: emergency department
ILI: Influenza-like Illness
MSS: Mortality Surveillance System
NCHS: National Center for Health Statistics
NSSP: National Syndromic Surveillance Program
NYC: New York City
PCR: Polymerase chain reaction
SARS-CoV-2: Severe Acute Respiratory Syndrome Coronavirus 2
US: United States

## Acknowledgements

We thank Farida Ahmen, Michael Augenbraun, Stephan Kohlhoff, Josh Marshall, and Tracey Wilson for helpful conversations, and our spouses/partners for childcare.

## Appendix Detailed methods

### Semiparametric excess mortality model

Covariates include week of the year, year, an indicator function that presents the possibility that the mean death rate has increased after the start of the emergency in a location, and the natural logarithm of population as an offset. Week of the year is modeled non- parametrically to capture seasonal mortality variations. Let *D*_*t*_ = number of certified deaths at time index *t, N*_*t*_ = population size. We use *p*_*t*_ as an indicator of time period *t* falls in. The indicator variable permit us to estimate excess deaths. Specifically, *p*_*t*_ = 0 represents the pre-emergency period; *p*_*t*_ = 1, the period after the emergency. Moreover, let *week*_*t*_ = week of year, and *year*_*t*_ = a categorical year effect.

Assuming *D*_*t*_ follows a Poisson distribution, we fit a semiparametric model;

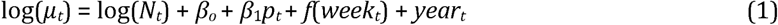

where *μ*_*t*_ = *E*(*D*_*t*_|*t,p*_*t*_,*N*_*t*_,week_*t*_,*year*_*t*_). The natural logarithm of *N*_*t*_ is an offset variable; while *f* is a smooth function of *week*, which accounts for within year variation. *f* is fit using a penalized cyclic cubic regression spline [19].

Preliminary analysis indicates overdispersion, so we used a quasi-Poisson model to estimate the dispersion parameter [17]. We estimated coefficients by a penalized likelihood maximization approach, where the smoothing penalty parameters are determined by restricted maximum likelihood. The residuals of the fitted model (1) did not present remaining temporal dependence.

The fit of model (1) can be used to estimate excess deaths through the difference between the estimated model with *p*_*t*_ = 1, versus the estimated model with *p*_*t*_ = 0. Let 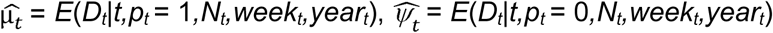, and 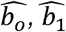, estimate *β_o_*, *β*_1_ respectively. Then [2],

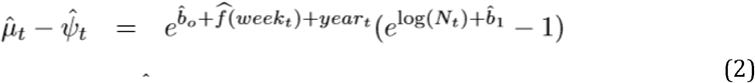

When *p*_*t*_ = 0, then 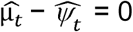. Equation (2) is the maximum likelihood estimator for expected excess deaths at *t*.

To estimate cumulative excess deaths we use (2),

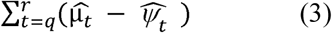

for any time period starting at index *q* and ending at *r*. Approximate simulations from the Bayesian posterior density are performed to obtain 95% credibility intervals, which we refer to as confidence intervals throughout.

### Conventional mortality method

Conventional excess mortality methods build a temporal mortality model until time *m < T*, then use this model to predict deaths from time *m* + 1 to time *T*, and excess deaths are the difference between the deaths between times *m* + 1 and *T*, and the predicted deaths from the model. Uncertainty is usually quantified through the uncertainty of the parameters on the regression model [16], which ignores natural random fluctuations in mortality from time *m* + 1 to time *T*. We tackle this problem combining the Bayesian posterior density described in section 3.3 with uncertainty on weekly mortality.

First, we fit a quasi-Poisson semiparametric model similar to the one presented in the previous section, but here we only use data until February 1, 2020. The deaths from February 8, 2020 forward should follow a Poisson distribution with some expected rate; the maximum likelihood estimator of such rate is the mean weekly deaths during this period. Theweekly deaths are approximately normal with a variance that accounts for overdispersion according to the scale parameter in the fitted model. From this distribution, we simulate 10,000 weekly deaths and subtract the results from the posterior distributions of the fitted results. The 95% confidence interval is the range between the 2.5% and 97.5% percentiles of all excess deaths.

## References

[1] André Ricardo Ribas Freitas, Maria Rita Donalisio, and Pedro María Alarcón-Elbal. Excess mortality and causes associated with chikungunya, puerto rico, 2014–2015. Emerging infectious diseases, 24(12):2352, 2018.

[2] Roberto Rivera and Wolfgang Rolke. Modeling excess deaths after a natural disaster with application to hurricane maria. Statistics in medicine, 38(23):4545–4554, 2019.

[3] Steven G. Krantz and Arni S. R. Srinivasa Rao. Level of underreporting including underdiagnosis before the first peak of COVID-19 in various countries: Preliminary retrospective results based on wavelets and deterministic modeling. Infection Control and Hospital Epidemiology, pages 1–3, April 2020.

[4] Wang Ma, Xiang Huo, and Minghao Zhou. The healthcare seeking rate of individuals with influenza like illness: A metaanalysis. Infectious Diseases, 50(10):728–735, October 2018. eprint: https://doi.org/10.1080/23744235.2018.1472805.

[5] Lone Simonsen, Peter Spreeuwenberg, Roger Lustig, Robert J Taylor, Douglas M Fleming, Madelon Kroneman, Maria D Van Kerkhove, Anthony W Mounts, W John Paget, et al. Global mortality estimates for the 2009 influenza pandemic from the glamor project: a modeling study. PLoS medicine, 10(11), 2013.

[6] Alyssa S Parpia, Martial L Ndeffo-Mbah, Natasha S Wenzel, and Alison P Galvani. Effects of response to 2014–2015 ebola outbreak on deaths from malaria, hiv/aids, and tuberculosis, West Africa. Emerging infectious diseases, 22(3):433, 2016.

[7] Sunil M. Shah, Iain M. Carey, Tess Harris, Stephen Dewilde, Christina R. Victor, and Derek G. Cook. The effect of unexpected bereavement on mortality in older couples. American Journal of Public Health, 103(6):1140–1145, June 2013.

[8] Nathan R. Hoot and Dominik Aronsky. Systematic Review of Emergency Department Crowding: Causes, Effects, and Solutions. Annals of Emergency Medicine, 52(2):126–136.e1, August 2008.

[9] Safiya Richardson, Jamie S. Hirsch, Mangala Narasimhan, James M. Crawford, Thomas McGinn, Karina W. Davidson, Douglas P. Barnaby, Lance B. Becker, John D. Chelico, Stuart L. Cohen, Jennifer Cookingham, Kevin Coppa, Michael A. Diefenbach, Andrew J. Dominello, Joan Duer-Hefele, Louise Falzon, Jordan Gitlin, Negin Hajizadeh, Tiffany G. Harvin, David A. Hirschwerk, Eun Ji Kim, Zachary M. Kozel, Lyndonna M. Marrast, Jazmin N. Mogavero, Gabrielle A. Osorio, Michael Qiu, and Theodoros P. Zanos. Presenting Characteristics, Comorbidities, and Outcomes Among 5700 Patients Hospitalized With COVID-19 in the New York City Area. JAMA, April 2020.

[10] Steffie Woolhandler and David U. Himmelstein. Intersecting U.S. Epidemics: COVID-19 and Lack of Health Insurance. Annals of Internal Medicine, April 2020.

[11] MR Spencer and F Ahmad. Timeliness of death certificate data for mortality surveillance and provisional estimates. vital statistics rapid release special report no. 001. us department of health and human services, centers for disease control and prevention. National Center for Health Statistics, National Vital Statistics System. https://www.cdc.gov/nchs/data/vsrr/report001.pdf, 2016.

[12] US Census Bureau. National Population by Characteristics: 2010-2019. https://www.census.gov/data/tables/time-series/demo/popest/2010snational-detail.html.

[13] Alexis Madrigal, Jeffrey Hammerbacher, Erin Kissane, and COVID Tracker team. COVID Tracking Project. https://covidtracking.com/data.

[14] New York Times. Coronavirus in the U.S.: Latest Map and Case Count. https://www.nytimes.com/interactive/2020/us/coronavirus-uscases.html, April 2020.

[15] CDC. Daily Updates of Totals by Week and State: Provisional Death Counts for Coronavirus Disease 2019 (COVID-19). https://www.cdc.gov/nchs/nvss/vsrr/COVID19/index.htm, 2020.

[16] Carlos Santos-Burgoa, John Sandberg, Erick Suárez, Ann Goldman-Hawes, Scott Zeger, Alejandra Garcia-Meza, Cynthia M Pérez, Noel Estrada Merly, Uriyoan Colón-Ramos, Cruz María Nazario, et al. Differential and persistent risk of excess mortality from hurricane Maria in Puerto Rico: a time-series analysis. The Lancet Planetary Health, 2(11):e478–e488, 2018.

[17] Jay M Ver Hoef and Peter L Boveng. Quasi-poisson vs. negative binomial regression: how should we model overdispersed count data?Ecology, 88(11):2766–2772, 2007.

[18] Quealy K. The Upshot: The Richest Neighborhoods emptied out most as coronavirus hit New York City [original data analysis]. New York Times. May 15, 2020..

[19] S. N. Wood. Generalized additive models: an introduction with R. Chapman and Hall/CRC, 2017.

[20] R Core Team. R: A Language and Environment for Statistical Computing. R Foundation for Statistical Computing, Vienna, Austria, 2016.

[21] National Vital Statistics System. Guidance for Certifying Deaths Due to Coronavirus Disease 2019 (COVID–19). National Vital Statistics System: Vital Statistics Reporting Guidance, (Report 3), April 2020.

[22] CDC. Coronavirus disease 2019 (COVID-19): Collection and submission of postmortem specimens from deceased persons with known or suspected COVID-19,March 2020 (interim guidance). https://www.cdc.gov/coronavirus/2019-ncov/hcp/guidance-postmortemspecimens.html, 2020.

[23] New York City Department of Health and Mental Hygiene. Influenza Surveillance Report Week ending April 18, 2020 (Week 16).

[24] CDC. COVIDView: A weekly surveillance summary of U.S. COVID-19 activity. https://www.cdc.gov/coronavirus/2019-ncov/coviddata/covidview/index.html, May 2020.

[25] M Boukhris, A Hillani, F Moroni, and et al. Cardiovascular implications of the covid-19 pandemic: A global perspective. Can J Cardiol., S0828282X(20):30464–5, 2020.

[26] CDC. Leading Causes of Death: Data are for the U.S. https://www.cdc.gov/nchs/fastats/leading-causes-of-death.htm, May 2020.

[27] Matthew Biggerstaff, Michael A. Jhung, Carrie Reed, Alicia M. Fry, Lina Balluz, and Lyn Finelli. Influenza-like illness, the time to seek healthcare, and influenza antiviral receipt during the 2010-2011 influenza season-United States. The Journal of Infectious Diseases, 210(4):535–544, August 2014.

[28] Mark B. Parshall, Richard M. Schwartzstein, Lewis Adams, Robert B. Banzett, Harold L. Manning, Jean Bourbeau, Peter M. Calverley, Audrey G. Gift, Andrew Harver, Suzanne C. Lareau, Donald A. Mahler, Paula M. Meek, and Denis E. O’Donnell. An Official American Thoracic Society Statement: Update on the Mechanisms, Assessment, and Management of Dyspnea. American Journal of Respiratory and Critical Care Medicine, 185(4):435–452, February 2012.

[29] Ruoqing Li, Jigang Tian, Fang Yang, Lei Lv, Jie Yu, Guangyan Sun, Yu Ma, Xiaojuan Yang, and Jianqiang Ding. Clinical characteristics of 225 patients with COVID-19 in a tertiary Hospital near Wuhan, China. Journal of Clinical Virology: The Official Publication of the Pan American Society for Clinical Virology, 127:104363, April 2020.

[30] Long-Quan Li, Tian Huang, Yong-Qing Wang, Zheng-Ping Wang, Yuan Liang, Tao-Bi Huang, Hui-Yun Zhang, Weiming Sun, and Yuping Wang. COVID-19 patients’ clinical characteristics, discharge rate, and fatality rate of meta-analysis. Journal of Medical Virology, March 2020.

[31] Francois-Xavier Lescure, Lila Bouadma, Duc Nguyen, Marion Parisey, Paul-Henri Wicky, Sylvie Behillil, Alexandre Gaymard, Maude Bouscambert-Duchamp, Flora Donati, Quentin [Le Hingrat, Vincent Enouf, Nadhira Houhou-Fidouh, Martine Valette, Alexandra Mailles, Jean-Christophe Lucet, France Mentre, Xavier Duval, Diane Descamps, Denis Malvy, Jean-François Timsit, Bruno Lina, Sylvie van-der-Werf, and Yazdan Yazdanpanah. Clinical and virological data of the first cases of COVID-19 in Europe: A case series. The Lancet Infectious Diseases, 2020.

[32] Ling Lin, Lianfeng Lu, Wei Cao, and Taisheng Li. Hypothesis for potential pathogenesis of SARS-CoV-2 infection-a review of immune changes in patients with viral pneumonia. Emerging Microbes & Infections, 9(1):727–732, December 2020.

[33] Tao Guo, Yongzhen Fan, Ming Chen, Xiaoyan Wu, Lin Zhang, Tao He, Hairong Wang, Jing Wan, Xinghuan Wang, and Zhibing Lu. Cardiovascular Implications of Fatal Outcomes of Patients With Coronavirus Disease 2019 (COVID-19). JAMA cardiology, March 2020.

[34] Yichun Cheng, Ran Luo, Kun Wang, Meng Zhang, Zhixiang Wang, Lei Dong, Junhua Li, Ying Yao, Shuwang Ge, and Gang Xu. Kidney disease is associated with in-hospital death of patients with COVID-19. Kidney International, 97(5):829–838, May 2020.

[35] Evangelos Terpos, Ioannis Ntanasis-Stathopoulos, Ismail Elalamy, Efstathios Kastritis, Theodoros N. Sergentanis, Marianna Politou, Theodora Psaltopoulou, Grigoris Gerotziafas, and Meletios A. Dimopoulos. Hematological findings and complications of COVID-19. American Journal of Hematology, April 2020.

[36] Ruppert David, Wand M. P., and Carroll R.J. (2003) Semiparametric Regression. Cambridge. 2003

[37] Theodore Lytras, Katerina Pantavou, Elisavet Mouratidou, and Sotirios Tsiodras. Mortality attributable to seasonal influenza in Greece, 2013 to 2017: Variation by type/subtype and age, and a possible harvesting effect. Euro Surveillance: Bulletin Europeen Sur Les Maladies Transmissibles = European Communicable Disease Bulletin, 24(14), April 2019.

[38] Claire Margerison-Zilko, Sidra Goldman-Mellor, April Falconi, and Janelle Downing. Health Impacts of the Great Recession: A Critical Review. Current epidemiology reports, 3(1):81–91, March 2016.

[39] Marc Lipsitch, David L. Swerdlow, and Lyn Finelli. Defining the Epidemiology of Covid-19 — Studies Needed. New England Journal of Medicine, 382(13):1194–1196, March 2020. eprint: https://doi.org/10.1056/NEJMp2002125

[40] Diana M. Prieto, Tapas K. Das, Alex A. Savachkin, Andres Uribe, Ricardo Izurieta, and Sharad Malavade. A systematic review to identify areas of enhancements of pandemic simulation models for operational use at provincial and local levels. BMC public health, 12:251, March 2012.

